# Use of health care services during the Covid-19 pandemic in Ethiopia: Evidence from a health facility survey

**DOI:** 10.1101/2021.08.09.21261754

**Authors:** Zemzem Shigute, Anagaw Mebratie, Getnet Alemu, Matthias Rieger, Arjun S. Bedi

## Abstract

**Introduction:** In recent years Ethiopia has made enormous strides in enhancing access to health care, especially, maternal and child health care (MCH). With the onset and spread of Covid-19, the attention of the health care system has pivoted to handling the disease, *potentially* at the cost of other health care needs. This paper explores whether this shift has come at the cost of non-Covid related health care, especially the use of MCH services.

**Methods:** Graphs, descriptive statistics and paired *t-tests* of significance are used to compare levels of inpatient and outpatient health care service utilization before and after the onset and spread of the virus. The analysis is based on a survey of 59 health centers and 29 public hospitals located in urban Ethiopia – the most acutely affected region of the country. Data on the use of health care services for a period of 24 months was gathered from the health management information systems (HMIS) of these facilities.

**Results:** There is a sharp reduction in the use of both inpatient (20-27%) and outpatient (27-34%) care, particularly in Addis Ababa, which has been most acutely affected by the virus. However, the decline does not come at the cost of MCH services. The use of several MCH components (skilled birth attendant deliveries, immunization, post-natal care) remains unaffected throughout the period while others (family planning services, ante-natal care) experience a decline (8-17%) in the immediate aftermath but recover soon after.

**Conclusion:** Concerns about the crowding out of MCH services due to the focus on Covid 19 are unfounded. Pro-active measures taken by the government and health care facilities to ring-fence the use of essential health care services have mitigated service disruptions. The results underline the resilience and agility displayed by one of the world’s most resource-constrained health care systems.

Summary Box

What is already known?

- Simulation studies suggest sharp increases in unwanted pregnancies, and in maternal and child mortality due to Covid related disruptions in the provision of mother and child health care services.
- Evidence on whether such service disruptions have indeed occurred is limited.

What are the new findings?

- Based on retrospective health facility data collected from parts of Ethiopia that have been most acutely affected by Covid, we find that for most MCH services, disruption is temporary and rebounds to pre-covid levels within two months.

What do the new findings imply?

- Concerns about the crowding out of MCH services due to the focus on Covid seem to be unfounded.
- Pro-active measures taken by the government and health care facilities to adapt and to ring-fence the use of essential health care services have mitigated service disruptions.
- Despite resource-constraints it is possible to protect the provision of essential health care services and thereby hard-won gains in maternal and child health outcomes

## Introduction

In the past 20 years, Ethiopia has witnessed a sharp expansion in its public healthcare system. Between 2000 and 2019 there has been a 21-fold increase in the number of health posts, an 11-fold increase in the number of health centers and a four-fold increase in the number of public hospitals [1-2]. Per capita healthcare spending has grown from US$5.4 in 2000 to US$24.2 in 2018 [3]. These interventions in infrastructure have been underpinned by two large community initiatives – the expansion of the system of health extension workers (HEW) and the spread of the so-called health development army (HDA) – community-based networks of women who identify and monitor pregnant women and focus on encouraging their members to use maternal health services.

Due to these efforts, access to essential healthcare services, as measured by the availability of health posts or health centers within a two-hour walking distance has increased from 51% in 2000 to 94% in 2016 [4-5]. In addition to access, the various interventions have been associated with increases in the use of mother and child health services and various health gains [6-10]. For instance, between 2000 and 2019 the proportion of women receiving antenatal care from a skilled provider at least once for their last birth increased from 27% to 74% while deliveries attended by skilled birth attendants (SBA) or in health facilities increased from 6% to 50% over the same duration. In terms of outcomes, while still higher than the global maternal mortality ratio (MMR) of 216, Ethiopia’s MMR of 412 per 100,000 live births, implies a 71% decline in maternal mortality over the period 2000 to 2019 [11].

These impressive gains have taken place in the context of a strengthening but still weak and under-resourced health care system. Compounding these issues, the onset and spread of the Covid-19 virus since March 2020 and the need to pivot and equip the health care system towards tackling the virus has led to concerns about the sustainability of these hard-won gains in maternal and child health outcomes [12-13]. Such concerns are not unique to Ethiopia. Disruptions to maternal and child health care services may translate into unintended pregnancies and increased maternal and child morbidity and mortality, especially in low-income countries. Based on simulations which allow for varying levels of disruption in access to MCH care and decreased access to food across 118 low- and middle-income countries, there are projections of more than 200,000 child deaths and 12,000 maternal deaths in the least severe scenario [14]. Similarly, based on the 37 countries, including Ethiopia, in which Marie Stopes International operates, there are projections of an additional 1.3 million unintended pregnancies and an additional 5,000 pregnancy-related deaths [15]. Based on telephone surveys, conducted during the period July to November 2020, in Addis Ababa, Ouagadougou and Lagos, 28-39% of health care providers perceived reductions in provision of child health care [16]. Consistent with such concerns there have been calls to ring-fence access to essential health services, especially to MCH care [17-22].

In Ethiopia, soon after the detection of the virus, the country went into a strict lockdown and the country’s hospitals, especially in the capital were re-purposed to treat virus affected patients with, *potentially*, adverse consequences for other health care needs. For instance, some public hospitals were designated as Covid only hospitals and stopped provision of outpatient and inpatient services for non-Covid health issues [12, 23]. Other concerns included, absenteeism of health care personnel due to lack of safety equipment [24]. On the demand-side, the fear of contracting the virus and travel restrictions is likely to have translated into reductions in the use of ante-natal and post-natal care and an increase in non-institutional births.

Whether dire predictions based on simulations [14, 15, 25] come to fruition in low-income countries such as Ethiopia or whether concerns such as health worker absenteeism, and reduced uptake of health services, which emerged soon after the imposition of virus-control measures are short-lived, is not yet clear. A search of the literature revealed no dearth of papers [17-22] calling for attempts to protect the provision of MCH in low-income countries, particularly in Sub-Saharan Africa (SSA) but evidence on pre- and post-pandemic use of health services in Ethiopia and more generally in SSA was more limited.

Comprehensive analyses of changes in health service utilization is provided in [26], who use country-wide HMIS data from 8 SSA countries (Cameroon, Democratic Republic of Congo, Liberia, Malawi, Mali, Nigeria, Sierra Leone and Somalia) to analyze the effect of Covid-19 on the use of MCH services. Their analysis shows that outpatient consultations and child vaccinations were the most vulnerable services while decreases in maternal health care were less generalized. For instance, between March and July 2020, the use of outpatient services declined from a low of 1% in Somalia to a high of 16% in Nigeria. Except for the DRC, vaccination shots (Pentavalent 3) fell between 2% in Cameroon to 17% in Mali. Although, not universal, the declines were most acute in April-May with a reversal in June-July. Single country analysis of changes in access to MCH, which perhaps allows for deeper exploration are also available [27, 28]. HMIS data from Rwanda is used to compare the use of MCH services between March-April 2019 and March-April 2020 - the authors report statistically significant declines in the use of 15 of 30 MCH services [27]. Analysis based on HMIS data from South Africa’s KwaZulu Natal province and a pre-post comparison yields sharp and significant declines for clinic attendance (36%) and hospital admissions (50%) for children aged less than 5 years in the period April-June 2020 [28].

There is also an emerging body of work in the Ethiopian context. A comparison of MCH use between March – July 2020 and March – July 2019 in 15 health centers in rural Ethiopia finds modest declines in key MCH and nutrition services which are pronounced in March and April and recover, thereafter [29]. However, since the main effect of the pandemic was in urban Ethiopia a focus on rural parts is likely to underestimate the consequences. Analysis of data gathered from 31 health centers and four hospitals located in SNNPR region displays reduction in the use of ANC, health facility births, PNC, family planning visits, and newborn immunization services with the effect ranging from 16 to 29 percent. There is evidence of an uptake in services in May and June 2020 after reductions in March and April 2020 (30). In terms of the incidence of Covid-19 cases, in August 2020, the region accounted for 1.6% of the cases in the country. Closest to the current contribution is the work reported in [30] who examine changes in health care use between November 2019 and June 2020 in nine health facilities located in Addis Ababa, the region most acutely affected by the virus. They report a 25% reduction in inpatient care and a 42% reduction in outpatient visits. In marked contrast the use of MCH services yields a more sanguine picture with the use of ANC1, ANC4, Penta1, Penta4 vaccinations and the use of skilled birth attendants (SBA) displaying no declines before and during COVID-19.

This paper contributes to the body of work on the effects of COVID-19 on actual use of health services in SSA and in particular in Ethiopia. Such evidence is clearly needed to identify vulnerabilities in the health care system and more importantly to support the continued provision of essential services to protect hard won gains in maternal and child mortality. This paper responds to such concerns and uses data culled from the HMIS of 88 health facilities located in four regional states of urban Ethiopia, as opposed to just a single region, to compare health care use in the four months (March-June 2020) immediately after the identification of the first case of the virus in the country with health care use a year before (March-June 2019) and the four months just preceding March 2020 (November 2019-February 2020). The paper examines changes in the use of both inpatient and outpatient care and thereafter focuses on the use of a range of MCH services. Since a key supply-side concern is the availability of health care services during the pandemic, we also examine health worker absenteeism before and after March 2020.

## Data and Methods

### Data

This study is based on a retrospective cross sectional health facility survey, conducted through phone and internet, which covered four regional states (Tigray, Amhara, Oromia and SNNP) and Addis Ababa city administration. Together, these regions account for 89.5 percent of the country’s population [31] and 85.6% of the Covid-19 cases in the country as per August 16, 2020 – the date that data collection commenced [32].

Ethiopia confirmed its first Covid-19 case on March 13, 2020 and data collection took place between August and December 2020. The plan was to cover 60 health centers and 30 public hospitals while the survey actually covered 59 health centers and 29 hospitals. The regional distribution of the sample was guided by the regional distribution of Covid-19 cases in the country at the time of the survey. Based on these considerations, the bulk of the sampled facilities were in Addis Ababa which accounted for 64% of Covid cases in August 2020, followed by Oromia (10%), Tigray (6%) and Amhara (4%) regions. There are 91 health centers in Addis Ababa and 44 were randomly chosen for the survey. The city has 12 public hospitals of which 11 were included in the survey (see Table 1). Since the sample is self-weighted towards those parts of the country that are most heavily affected by the virus, the results may be thought of as an upper bound in terms of the effect of Covid 19 on access to health services.

**Table 1.** Share of Covid Cases and Sample Distribution.

| Region | Covid-19 Cases<br>August 16, 2020 | Share of<br>region in<br>total cases<br>(%) | Health centers | Public<br>hospitals | Total |
| --- | --- | --- | --- | --- | --- |
|  | *N* |  |  |  |  |
| Addis Ababa | 19,188 | 64.2 | 44 | 11 | 55 |
| Oromia | 2,965 | 9.92 | 7 | 8 | 15 |
| Tigray | 1,772 | 5.93 | 3 | 5 | 8 |
| Amhara | 1,193 | 3.99 | 3 | 3 | 6 |
| SNNP | 477 | 1.59 | 2 | 2 | 4 |
| *Total* | *25,595* | *85.6* | *59* | *29* | *88* |

**Source:** Covid figures are from the National Public Health Emergency Operation Center (PHEOC) COVID-19 Weekly bulletin 16. The total number of confirmed cases in the country on August 16, 2020 was 29,876.

Based on prior discussions and consent, a survey instrument was sent by e-mail to health facility ICT workers responsible for their facilities’ health management information systems (HMIS). To ensure data quality, several rounds of iterative discussions took place with the ICT workers throughout the data collection and data cleaning process. From the HMIS, information was culled on monthly inpatient and outpatient visits for 24 months (July 2018 till June 2020). In addition to the total visits, data were collected on specific outpatient services including the gamut of maternal and childcare health services (family planning services (FPS), ante-natal care (ANC), abortions, delivery, post-natal care (PNC), immunization, integrated management of neonatal and childhood illnesses (IMNCI), prevention of mother-to-child HIV transmission care (PMTCT).

The survey also included modules to ascertain the level of preparedness and provision of Covid-19 related services, provision of supplies such as personal protective equipment, measures taken to mitigate/prevent the spread of Covid-19 and the challenges faced by the hospital due to the virus, especially related to absenteeism of health care professionals and support staff.

### Methods

Graphs, descriptive statistics and paired *t-tests* of significance are used to compare levels of inpatient and outpatient health care service utilization (number of patients per month) before and after the onset and spread of the virus. The analysis of outpatient visits includes data from health centers and public hospitals while inpatient care utilization is restricted to data from public hospitals as health centers mainly provide out-patient services. In addition to the total visits for health care, we also compare the number of visits for different types of mother and child health services such as family planning, antenatal care, abortion, delivery, postnatal care, immunization, IMNCI, PMTCT. We assess absenteeism of hospital staff by comparing the number of staff members that should be in the hospital and the number of staff members that were present on the day of the survey.

Although we have two years of utilization data, the focus is on the 4-month period, March to June 2020 as compared to health care use in the period March to June 2019. Comparing service use volumes for similar months, as opposed to before and after March 2020, helps to account for seasonal variations in patient flows (33). However, to account for the possibility that year-on-year comparisons may underestimate the extent of the decline, if there is an increase in health care utilization over time, we also compare health care use between March-June 2020 with a 4-month period (November 2019-February 2020), just preceding the first Covid case in Ethiopia. Graphs are used to demonstrate year-on-year changes in health care use, that is, comparing health care use, per month, over the period July 2019-June 2020 to the period July 2018-June 2019.

### Ethics clearance

Ethics approval (Ethics 2020-24) was provided by the Research Ethics Committee of the International Institute of Social Studies, Erasmus University Rotterdam.

### Public involvement

The research questions were developed based on conversations with and concerns about declines in non-Covid health care use and health worker absenteeism expressed by health care providers in the country’s largest hospital located in Addis Ababa. Suggestions on appropriate comparison periods and the type of health care services to be investigated were provided by managers of health care facilities. The results have been discussed with and corroborated by health care workers at two large hospitals in Addis Ababa.

## Results

Our discussion of the results begins by examining changes in the use of in-patient care, followed by commentary on out-patient care and its various components.

### Inpatient care use

Changes in the use of inpatient care are displayed in Table 2 and Figure 1. On average, per month, between March-June 2020, there were 558 inpatient admissions to the 29 hospitals. This is a 20% decline as compared to the 697 admissions a year ago and a 27% decline as compared to the 4-month period preceding March 2020. Regardless of the comparison period the reduction is statistically significant at the 1% significance level. Based on a year-on-year comparison, the declines are sharper in the regions most affected by the virus, that is, Addis Ababa and Oromiya which experience declines of about 24 - 26% as compared to other parts of the country which experience smaller (16% in Tigray; 9.5% in Amhara) or no declines (SNNPR). Year-on-year monthly comparisons (Figure 1) over the period show a sharp 40% decline in inpatient admissions between April 2020 and April 2019 and a similar decline. While this is a large decline, there are clear signs of recovery in May and June 2020 with utilization rates at about 80% of what they were in May-June 2019.

**Table 2.** Inpatient and Outpatient Health Care Use.

| **Average Monthly Inpatient Admissions in Public Hospitals** |  |  |  |  |  |  |
| --- | --- | --- | --- | --- | --- | --- |
| Region | *N* | March – June<br>2020<br>(1) | March – June<br>2019<br>(2) | Nov. 2019<br>– Feb. 2020<br>(3) | P-value<br>*Ho*: (1=2) | P-value<br>*Ho*: (1=3) |
| Addis Ababa | 11 | 656 | 861 | 910 | 0.022 | 0.007 |
| Oromia | 8 | 457 | 613 | 736 | 0.077 | 0.034 |
| Tigray | 5 | 313 | 371 | 409 | 0.319 | 0.236 |
| Amhara | 3 | 569 | 629 | 692 | 0.418 | 0.251 |
| SNNP | 2 | 1016 | 1042 | 1056 | 0.794 | 0.754 |
| *Total* | *29* | *558* | *697* | *763* | *0.001* | *0.000* |
| **Average Monthly Outpatient Visits to Health Centers and Public Hospitals** |  |  |  |  |  |  |
| Addis Ababa | 55 | 4318 | 6636 | 7038 | 0.004 | 0.000 |
| Oromia | 15 | 3461 | 4010 | 5082 | 0.235 | 0.045 |
| Tigray | 8 | 9262 | 11942 | 12254 | 0.065 | 0.014 |
| Amhara | 6 | 7008 | 8059 | 8926 | 0.408 | 0.194 |
| SNNP | 4 | 5903 | 5477 | 7334 | 0.606 | 0.214 |
| *Total* | *88* | *4877* | *6715* | *7321* | *0.001* | *0.000* |

**Figure 1:**
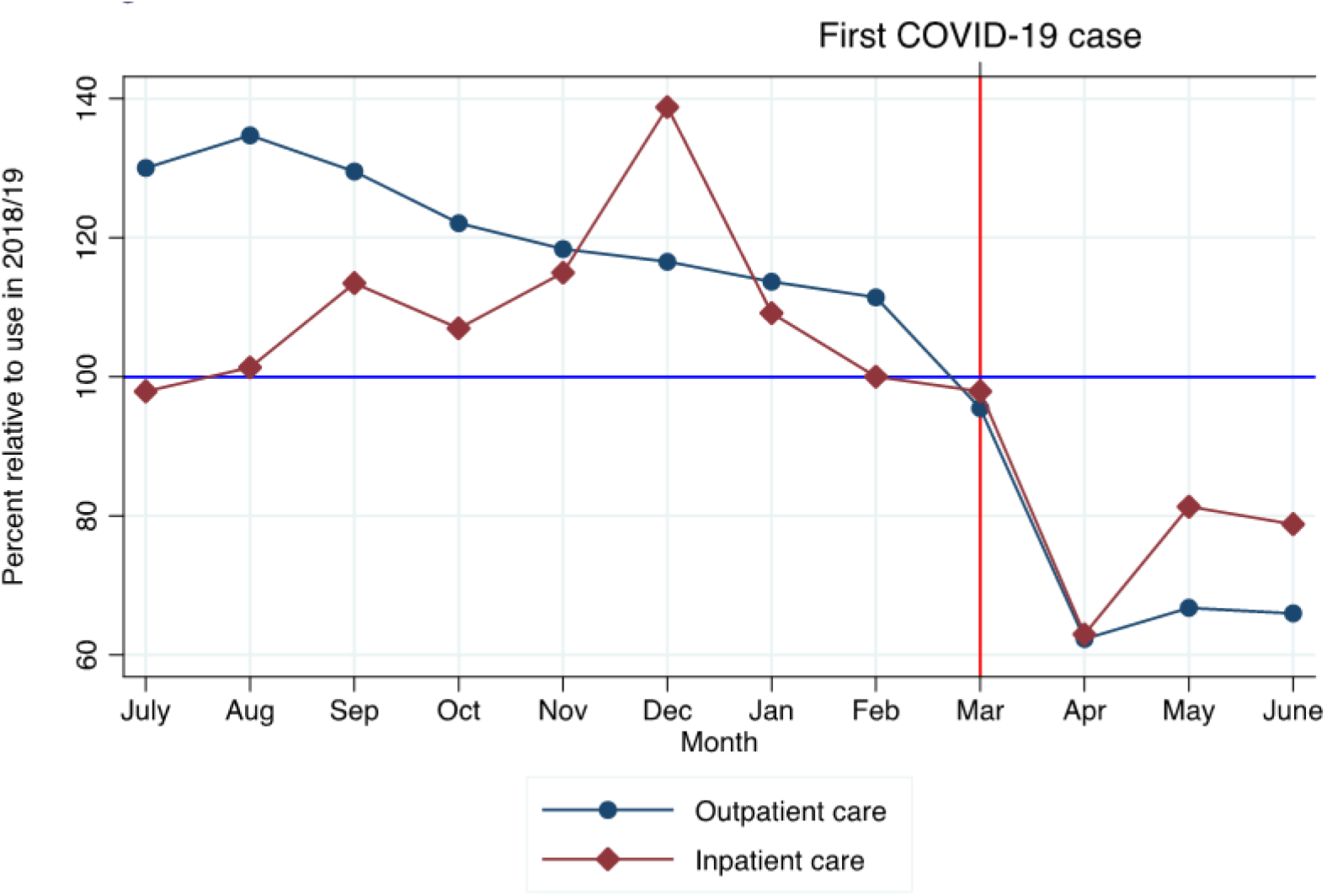
Volume of healthcare use in 2019/20 relative to same month in 2018/19

### Outpatient care use

The use of outpatient care is likely to be more discretionary as compared to admission for inpatient treatment and more likely to experience a decline. This does seem to be the case (Table 2), as the average monthly outpatient visits to sampled facilities between March and June 2020 was 4,877 while the corresponding figure between March - June 2019 was 6,715 or a decline of 27%. As compared to the four months immediately preceding March 2020, the decline in usage is even more pronounced (34%). Based on a year-on-year comparison the decline in Addis Ababa is 35% while it is about 39% based on comparing usage rates just prior to March 2020. As in the case of inpatient services the decline is pronounced in Addis Ababa as compared to other regions which experience smaller declines (14%, 22% and 13% in Oromia, Tigray and Amhara, respectively) or even a modest increase (SNNPR). Year-on-year monthly comparisons (Figure 1) shows a sharp 40% decline in health care use between March and April 2020 which begins to pick up in May and June 2020 but unlike inpatient care the uptick is much lower and throughout May and June 2020 the number of outpatient visits remains at about 65% of the pre-pandemic period.

### Utilization by type of service - Pre-Birth and Delivery

Given the decline in the use of both outpatient and inpatient care health use a key concern is whether usage of MCH services has been affected. Table 3 examines changes in the use of family planning services, antenatal care, abortions and deliveries.

**Table 3.** Mother and Child Health Care Use - Pre-Birth and Delivery Average Monthly Visits.

| **Family Planning Services** |  |  |  |  |  |  |
| --- | --- | --- | --- | --- | --- | --- |
| Region | *N* | March – June<br>2020<br>(1) | March – June<br>2019<br>(2) | Nov. 2019<br>– Feb. 2020<br>(3) | P-value<br>*Ho*: (1=2) | P-value<br>*Ho*: (1=3) |
| Addis Ababa | 54 | 63 | 71 | 73 | 0.184 | 0.002 |
| Oromia | 15 | 66 | 56 | 69 | 0.258 | 0.213 |
| Tigray | 8 | 93 | 125 | 131 | 0.199 | 0.084 |
| Amhara | 6 | 149 | 158 | 154 | 0.478 | 0.677 |
| SNNP | 4 | 76 | 71 | 107 | 0.776 | 0.436 |
| **Total** | **87** | **73** | **79** | **85** | **0.164** | **0.000** |
| **Antenatal Care** |  |  |  |  |  |  |
| Addis Ababa | 55 | 170 | 175 | 185 | 0.400 | 0.034 |
| Oromia | 15 | 145 | 162 | 174 | 0.167 | 0.058 |
| Tigray | 8 | 207 | 307 | 340 | 0.137 | 0.014 |
| Amhara | 6 | 153 | 185 | 196 | 0.282 | 0.019 |
| SNNP | 4 | 177 | 162 | 151 | 0.708 | 0.654 |
| **Total** | **88** | **168** | **185** | **197** | **0.031** | **0.000** |
| **Abortion care** |  |  |  |  |  |  |
| Addis Ababa | 53 | 13 | 16 | 17 | 0.026 | 0.002 |
| Oromia | 14 | 12 | 17 | 18 | 0.046 | 0.051 |
| Tigray | 8 | 22 | 27 | 34 | 0.491 | 0.067 |
| Amhara | 5 | 24 | 27 | 26 | 0.524 | 0.350 |
| SNNP | 4 | 19 | 22 | 27 | 0.465 | 0.153 |
| **Total** | **84** | **15** | **18** | **20** | **0.002** | **0.000** |
| **Delivery** |  |  |  |  |  |  |
| Addis Ababa | 50 | 145 | 135 | 129 | 0.130 | 0.073 |
| Oromia | 15 | 160 | 154 | 150 | 0.521 | 0.233 |
| Tigray | 7 | 197 | 192 | 189 | 0.567 | 0.021 |
| Amhara | 6 | 195 | 185 | 187 | 0.021 | 0.242 |
| SNNP | 3 | 164 | 178 | 215 | 0.094 | 0.171 |
| **Total** | **81** | **157** | **148** | **146** | **0.075** | **0.007** |

There is a small (8%) but statistically insignificant decline in the use of family planning services between March-June 2020 and March-June 2019. There is a more pronounced 16% decline in the use of family planning services between March-June 2020 and the period immediately preceding the onset of Covid in the country. There is a similar pattern with regard to the use of antenatal services. That is, a small (9%) decline based on a year-on-year comparison while the decline based on a comparison with the preceding period is 17% and statistically significant. Although the numbers are small in absolute terms, the use of health facilities for abortion care declines by about 16% based on a year-on-year comparison and by 25% as compared to the period November 2019 - February 2020. With regard to deliveries there are no changes over time, if anything, post-March 2020, there is a slight increase in the number of deliveries taking place at health facilities. Decomposing the overall change into monthly comparisons shows that declines in the use of FPS and ANC reach their nadir in March and April but recover to pre-pandemic levels in May and June. In contrast, the use of health services for abortions continues to decline post-March 2020 (Figures 2a, 2b).

**Figure 2a:**
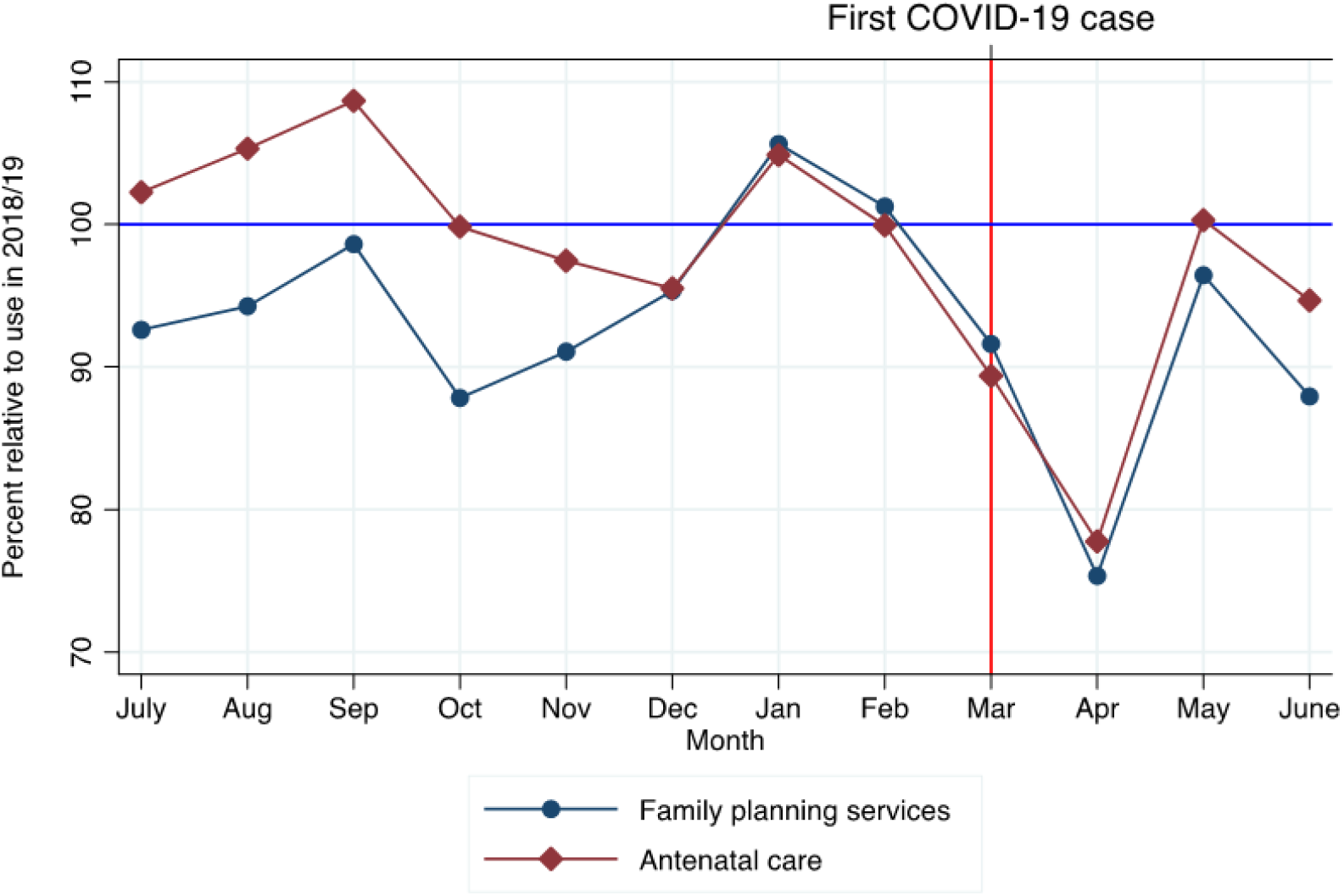
Volume of family planning service and ANC visitis in 2019/20 relative to same month in 2018/19

**Figure 2b:**
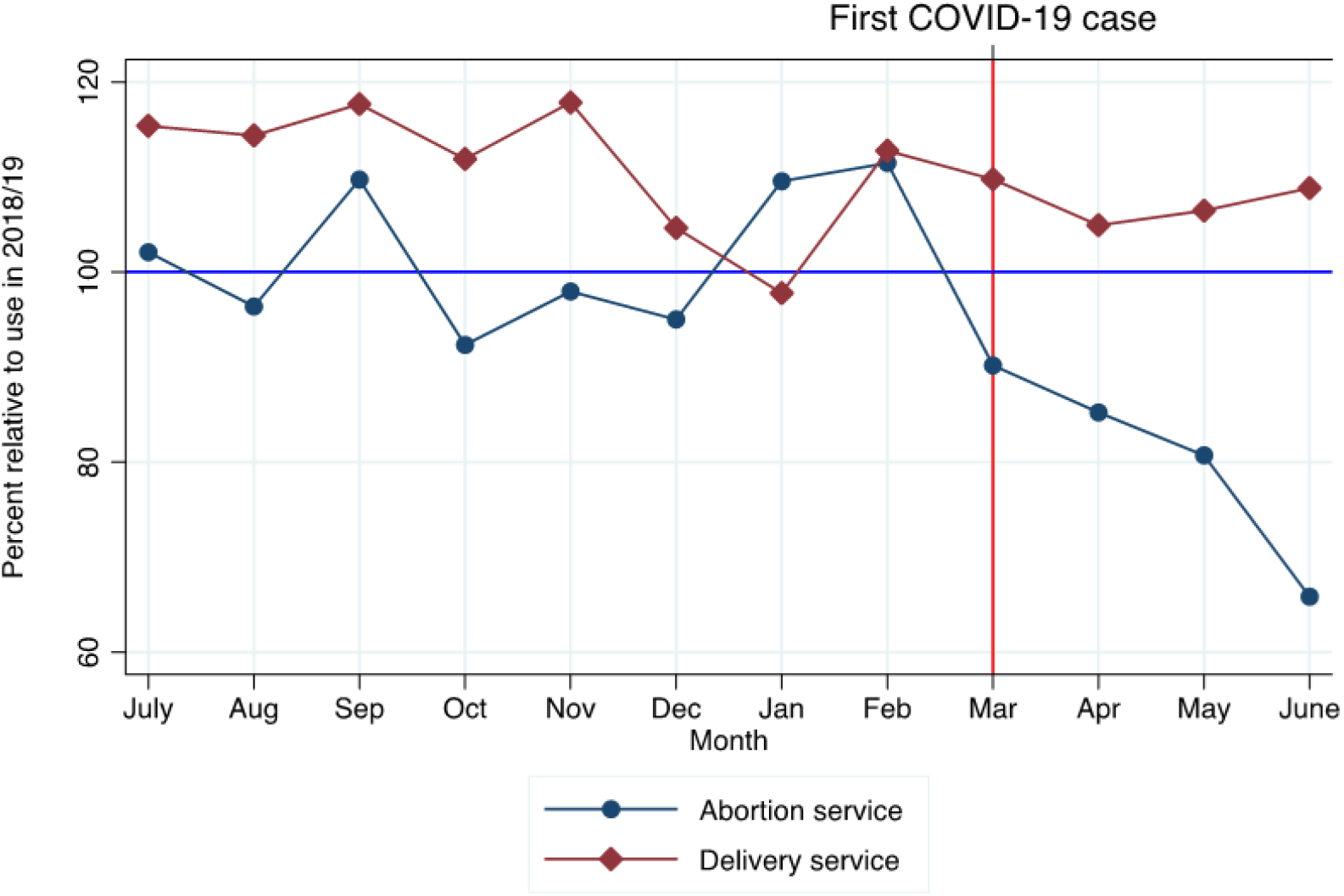
Volume of abortion and delivery service use in 2019/20 relative to same month in2018/19

### Utilization by type of service – Post-birth

The use of health facilities for postnatal care remains stable regardless of the reference period used for comparison (Table 4). Similarly, the use of immunization services does not seem to have experienced a major disruption. There is no change on the basis of year-on-year comparisons while there is a slight decline (6%) as compared to usage during the November 19-February 20 period with Addis Ababa experiencing the largest decline (10%). The disruption is largest in March 2020 but it is short-lived and by May-June 2020 the use of such services recovers to pre-pandemic levels. While postnatal care and immunization appear to be protected, there is a decline in the use of health services for PMTCT care of between 16-18% depending on the reference period under consideration. The effects are even larger for IMNCI care which experiences reductions of 16-29%, depending on the reference period. However, there are signs of recovery in May and June 2020, albeit use rates remain at about 90% of pre-pandemic levels (Figures 3a, 3b).

**Table 4.** Mother and Child Health Care Use – Post-birth Average Monthly Visits.

| **Postnatal care** |  |  |  |  |  |  |
| --- | --- | --- | --- | --- | --- | --- |
| Region | *N* | March – June<br>2020<br>(1) | March – June<br>2019<br>(2) | Nov. 2019<br>– Feb. 2020<br>(3) | P-value<br>*Ho*: (1=2) | P-value<br>*Ho*: (1=3) |
| Addis Ababa | 51 | 97 | 92 | 107 | 0.501 | 0.078 |
| Oromia | 15 | 169 | 150 | 161 | 0.340 | 0.690 |
| Tigray | 6 | 92 | 77 | 96 | 0.500 | 0.827 |
| Amhara | 6 | 85 | 75 | 99 | 0.290 | 0.200 |
| SNNP | 4 | 184 | 171 | 149 | 0.531 | 0.263 |
| **Total** | **82** | **113** | **104** | **118** | **0.138** | **0.323** |
| **Immunization** |  |  |  |  |  |  |
| Addis Ababa | 52 | 598 | 576 | 639 | 0.442 | 0.037 |
| Oromia | 15 | 592 | 714 | 617 | 0.267 | 0.513 |
| Tigray | 6 | 1376 | 1415 | 1591 | 0.873 | 0.171 |
| Amhara | 6 | 656 | 501 | 583 | 0.194 | 0.377 |
| SNNP | 4 | 664 | 643 | 732 | 0.773 | 0.357 |
| **Total** | **83** | **661** | **660** | **704** | **0.978** | **0.021** |
| **Prevention of mother-to-child HIV transmission (PMTCT) care** |  |  |  |  |  |  |
| Addis Ababa | 51 | 69 | 85 | 87 | 0.005 | 0.001 |
| Oromia | 11 | 119 | 131 | 122 | 0.567 | 0.218 |
| Tigray | 8 | 93 | 133 | 140 | 0.090 | 0.040 |
| Amhara | 6 | 133 | 145 | 168 | 0.606 | 0.161 |
| SNNP | 4 | 168 | 179 | 167 | 0.704 | 0.976 |
| **Total** | **80** | **88** | **105** | **107** | **0.002** | **0.000** |
| **Integrated Management of Neonatal and Childhood Illness (IMNCI) care** |  |  |  |  |  |  |
| Addis Ababa | 7 | 273 | 462 | 525 | 0.027 | 0.048 |
| Oromia | 6 | 205 | 229 | 191 | 0.272 | 0.807 |
| Tigray | 8 | 309 | 242 | 444 | 0.404 | 0.034 |
| Amhara | 5 | 254 | 326 | 310 | 0.454 | 0.067 |
| SNNP | 2 | 49 | 71 | 63 | 0.068 | 0.061 |
| **Total** | **28** | **249** | **297** | **353** | **0.182** | **0.002** |

**Figure 3a:**
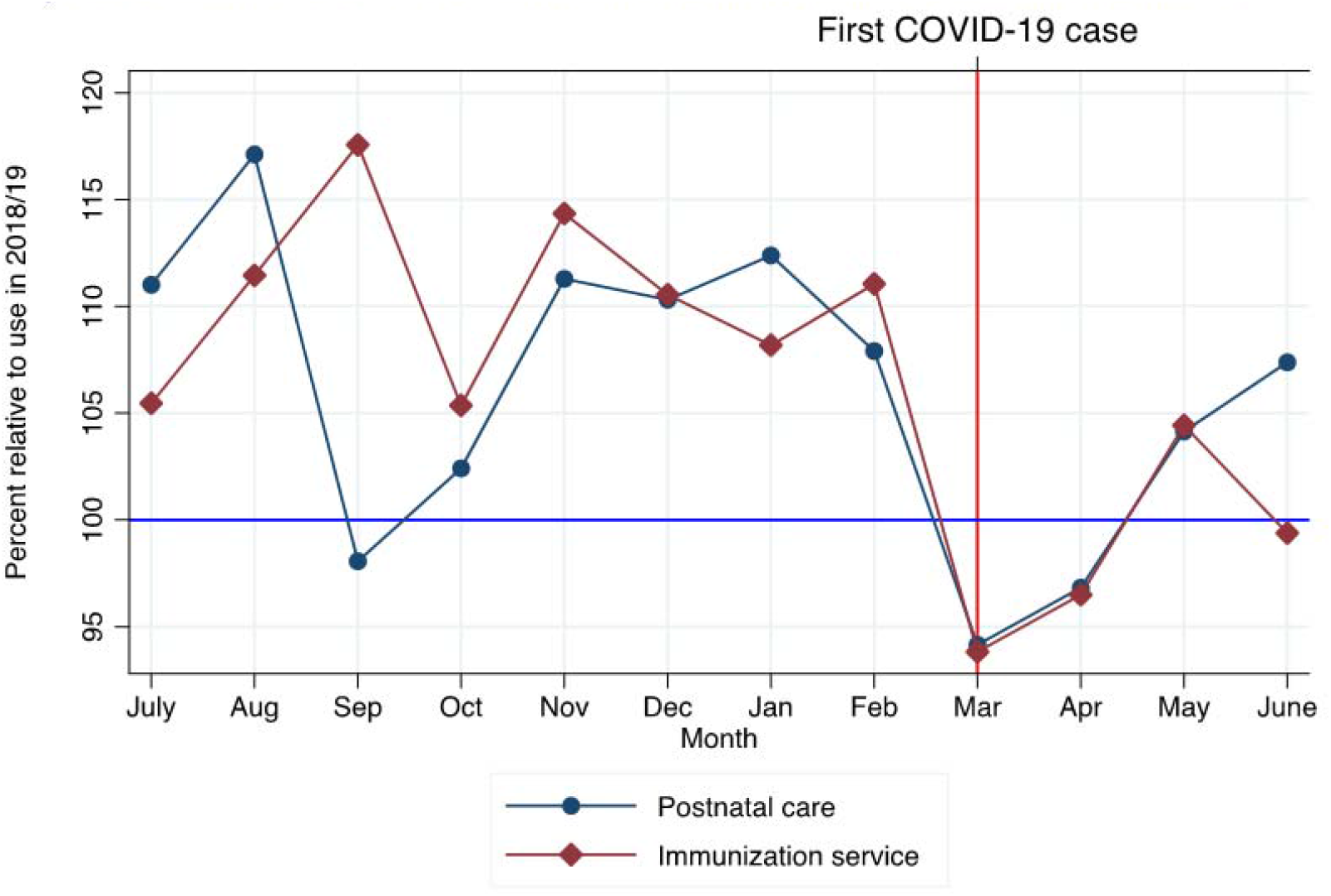
Volume of PNC and immunization service in 2019/20 relative to same month in 2018/19

**Figure 3b:**
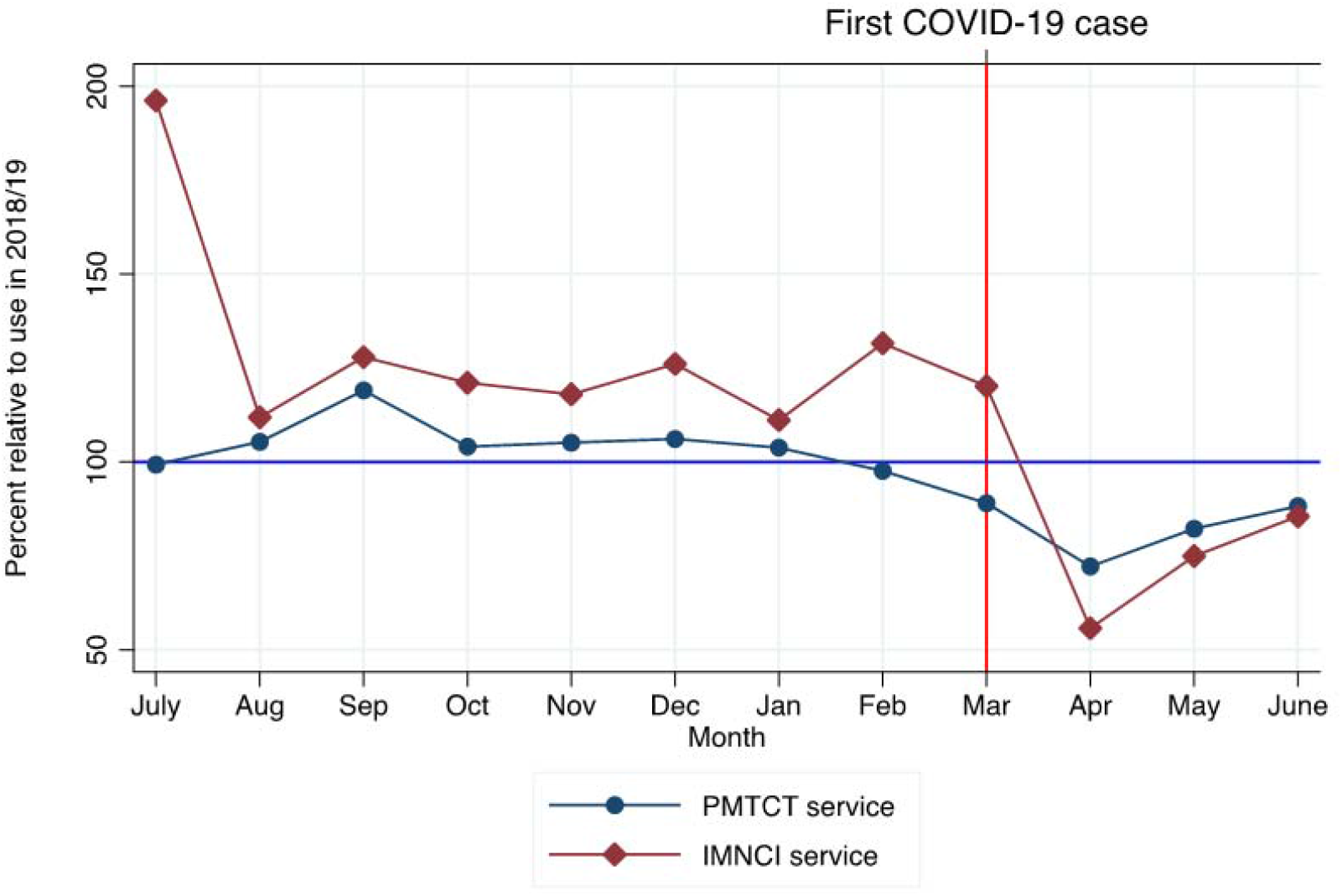
Volume of PMTCT and IMNCI serivices in 2019/20 relaive to same month in 2018/19

### Utilization of non-MCH health services

Table 2 and Figure 1 show clear declines in the use of health care services for both inpatient and outpatient care. Analysis of the different components of care shows that the decline in MCH services is not very pronounced, suggesting that the decline in health care usage comes from foregoing, postponing or substituting in-person hospital visits with alternative forms of care. While we do not have detailed information on all components of non-MCH care, the data we do have reveals a clear pattern. The data show that the use of health facilities for essential (ART, TB services) non-MCH care continues unabated (see Table 5), however, dental services and minor operating procedures show sharp declines. The use of health facilities for dental care drop by 45% and for minor operations by 35% on a year-on-year basis and shows very little recovery over time (see Figure 4).

**Table 5.**
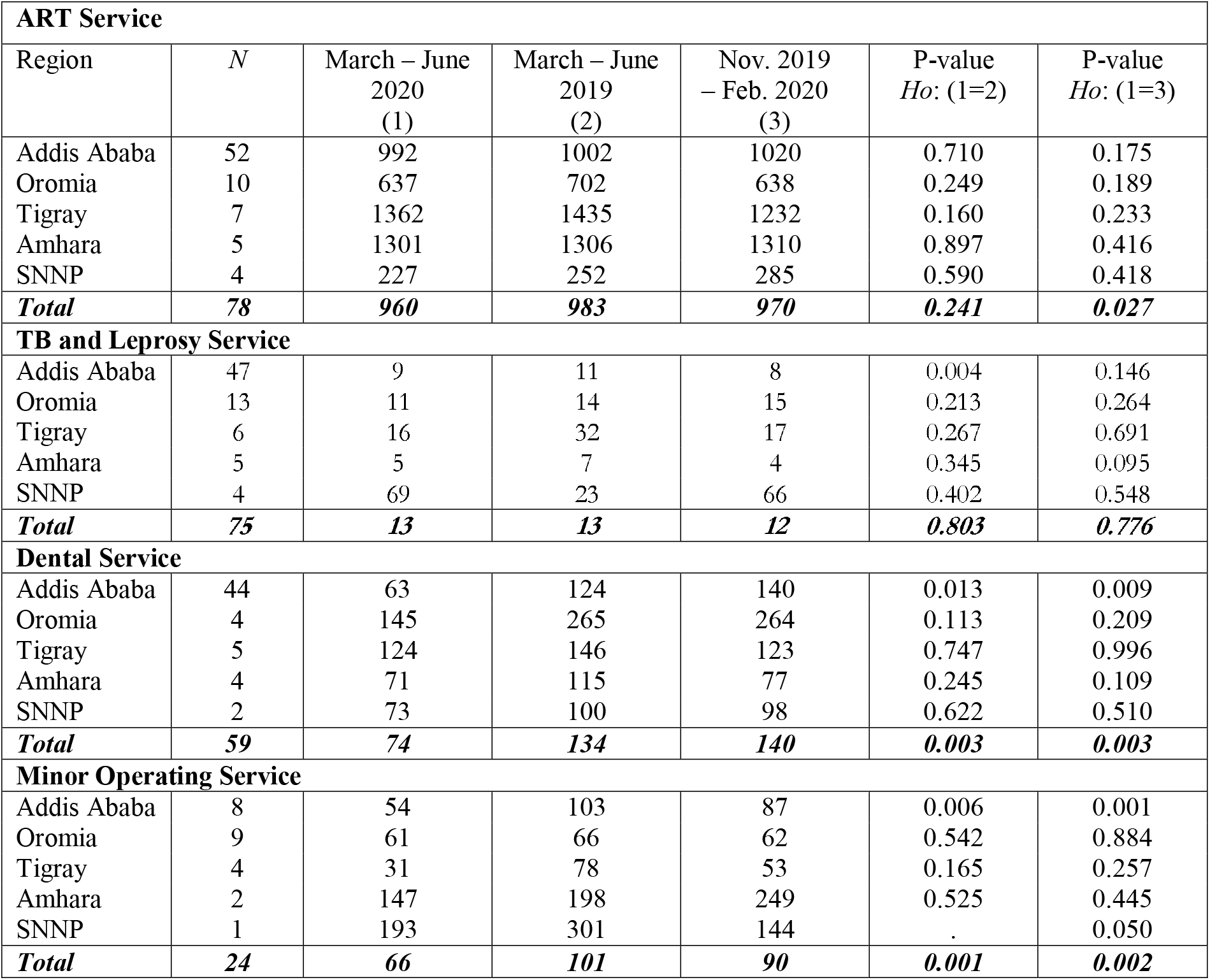
Utilization of various health services by adults Average Monthly Visits.

| **ART Service** |  |  |  |  |  |  |
| --- | --- | --- | --- | --- | --- | --- |
| Region | *N* | March – June<br>2020<br>(1) | March – June<br>2019<br>(2) | Nov. 2019<br>– Feb. 2020<br>(3) | P-value<br>*Ho*: (1=2) | P-value<br>*Ho*: (1=3) |
| Addis Ababa | 52 | 992 | 1002 | 1020 | 0.710 | 0.175 |
| Oromia | 10 | 637 | 702 | 638 | 0.249 | 0.189 |
| Tigray | 7 | 1362 | 1435 | 1232 | 0.160 | 0.233 |
| Amhara | 5 | 1301 | 1306 | 1310 | 0.897 | 0.416 |
| SNNP | 4 | 227 | 252 | 285 | 0.590 | 0.418 |
| **Total** | **78** | **960** | **983** | **970** | **0.241** | **0.027** |
| **TB and Leprosy Service** |  |  |  |  |  |  |
| Addis Ababa | 47 | 9 | 11 | 8 | 0.004 | 0.146 |
| Oromia | 13 | 11 | 14 | 15 | 0.213 | 0.264 |
| Tigray | 6 | 16 | 32 | 17 | 0.267 | 0.691 |
| Amhara | 5 | 5 | 7 | 4 | 0.345 | 0.095 |
| SNNP | 4 | 69 | 23 | 66 | 0.402 | 0.548 |
| **Total** | **75** | **13** | **13** | **12** | **0.803** | **0.776** |
| **Dental Service** |  |  |  |  |  |  |
| Addis Ababa | 44 | 63 | 124 | 140 | 0.013 | 0.009 |
| Oromia | 4 | 145 | 265 | 264 | 0.113 | 0.209 |
| Tigray | 5 | 124 | 146 | 123 | 0.747 | 0.996 |
| Amhara | 4 | 71 | 115 | 77 | 0.245 | 0.109 |
| SNNP | 2 | 73 | 100 | 98 | 0.622 | 0.510 |
| **Total** | **59** | **74** | **134** | **140** | **0.003** | **0.003** |
| **Minor Operating Service** |  |  |  |  |  |  |
| Addis Ababa | 8 | 54 | 103 | 87 | 0.006 | 0.001 |
| Oromia | 9 | 61 | 66 | 62 | 0.542 | 0.884 |
| Tigray | 4 | 31 | 78 | 53 | 0.165 | 0.257 |
| Amhara | 2 | 147 | 198 | 249 | 0.525 | 0.445 |
| SNNP | 1 | 193 | 301 | 144 | . | 0.050 |
| **Total** | **24** | **66** | **101** | **90** | **0.001** | **0.002** |

**Figure 4:**
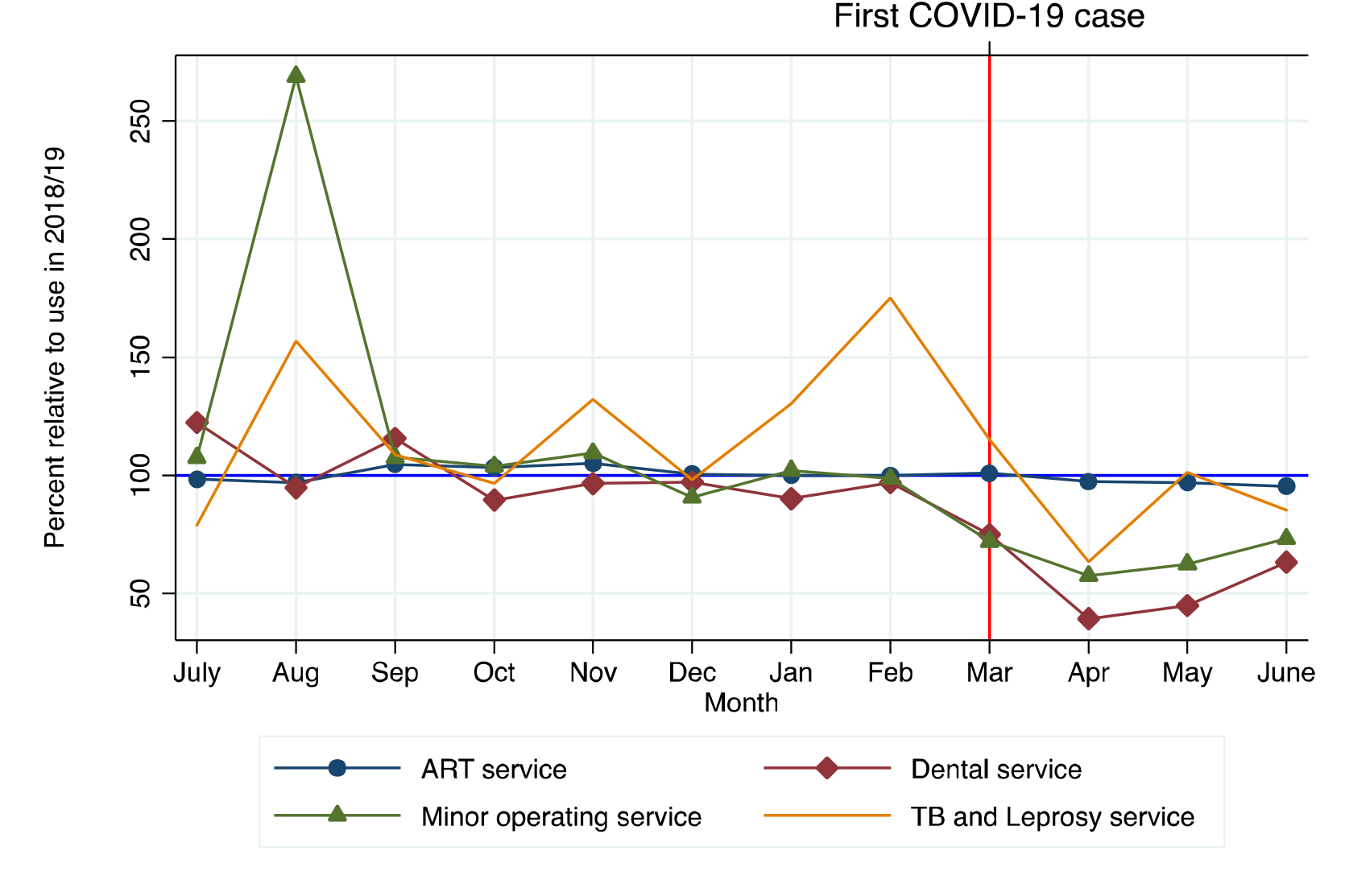
Volume of various health services in 2019/20 relative to same month in 2018/19

## Discussion

There is a clear and sharp decline in the use of both inpatient and outpatient services after the onset of Covid-19 in Ethiopia. Depending on the reference period the decline in inpatient care ranged between 20-27%. The decline in outpatient care is somewhat higher and ranges between 27-34%. The magnitude of the declines is similar to those reported in [23]. However, substantially higher as compared to the range of African countries analyzed in [26]. This is perhaps not surprising as the sample was designed to represent the most acutely affected regions of the country, with a majority of the sample coming from Addis Ababa and a majority of the health facilities in the sample (about 60%, see Table A1) are providing care for Covid infected patients. Thus, the estimates presented here should be viewed as an upper bound of the effect of Covid-19 on access to healthcare services in the country.

Unlike the sharp declines for overall use of health services, declines for several components of MCH care are lower and recovery is faster. For instance, with regard to MCH services, the use of FPS and ANC decline between 8-18% but there is no change in the use of health care facilities for deliveries, PNC and for immunization. These patterns are consistent with other work on Ethiopia [23, 29]. While considerable effort was made to check the completeness of the data obtained from the HMIS, health facility records may not always be complete. However, the consistency of the results presented here as compared to other work on Ethiopia [23,29] enhances the credibility of the HMIS data. Taken together the results support the idea that despite the sharp decline in overall use of health services, even in the most acutely affected parts of the country, Ethiopia has been able to protect the use of a number of essential maternal and child health care services. Recognition of the importance of maintaining “important patient care” has been articulated in the country’s Covid-19 management handbook [34] and by its political leadership [35]. Through mass media, consistent efforts have been made to communicate that public health facilities are prepared and are able to offer a safe environment for continued and safe use of health services [35].

The pattern of a decline followed by a return to pre-pandemic levels of MCH use in May and June 2020, highlights, both, the resilience and responsiveness of the health care system and the increase in confidence amongst health care users. Guidelines on preparing health facilities to deal with provision of services during the pandemic were rapidly established [34], infection prevention measures were systematically implemented by health facilities [29] and the delivery mode for some services was altered (from indoor buildings to outdoor tents; more frequent delivery of services in smaller groups). Increased provision and use of personal protective equipment – a key concern driving health worker absenteeism – was rapidly achieved. Our data show that all health facilities received additional equipment such as thermometers, face masks, and gloves, while about 70% received full PPE kits (suits, boots, face shades, goggles). Consistent with guidelines, almost all the facilities in our sample introduced a range of infection prevention measures (see Table A2). While health worker absenteeism seems to have increased during this time period (see Table A3), it is not particularly high in absolute terms (about 7%). Based on direct questions to facility managers, absenteeism of health worker staff was rated as an issue in the case of about 8% of the facilities in our sample. While there were several reasons for absenteeism the most prominent were that staff were on sanctioned leave, were on Covid duty elsewhere or were unwell, including due to the virus.

A recent brief [36] reviews Ethiopia’s policies to maintain essential health services in the time of Covid and argues that the approach does not call for substantive changes but is based on adaptations designed to strengthen the health system. The brief goes on to argue that “there is limited evidence on the relative effectiveness of this approach to maintain services during COVID-19”. The results presented in this paper along with the existing work on Ethiopia suggests that the approach has been successful.

## Conclusion

Simulation-based studies [14, 25] have provided disconcerting figures of the negative consequences of the disruption of MCH services and increased food insecurity on maternal and child mortality. At least from the perspective of health service disruptions, the results presented here along with those from other parts of Ethiopia and for a set of other countries in SSA (26) yield a more optimistic picture. However, these patterns are by no mean universal [27, 28]. In the case of Ethiopia, warnings to protect essential health care services, especially MCH seem to have been heeded and despite resource constraints, the country has successfully adapted and managed to protect the provision of essential health care services. While this is promising, there are MCH services such as PMTCT and IMNCI which have seen more sustained declines. Furthermore, there is no space for complacency as vaccination rates remain low and the virus continues to spread in rural parts of Ethiopia and SSA. Sustained efforts at maintaining balanced health care provision need to continue.

## Data Availability

The complete data set used for this study will be uploaded in a public repository upon acceptance.

## Acknowledgements

This work was supported by the D.P. Hoijer Fonds, Erasmus Trustfonds, Erasmus University Rotterdam, The Netherlands.

## Contributor statement

**ZS** Conceptualization, Survey design, Writing - Original Draft, Project Administration, Funding acquisition; **AM** Data collection, Data curation, Conceptualization, Methodology, Formal Analysis, Investigation, Writing - Original Draft; **GA** Data collection, Survey design, Conceptualization, Writing - Review & Editing; **MR** Methodology, Writing – Review and Editing, Funding acquisition; **AB** Conceptualization, Funding acquisition, Writing – Original Draft

All authors confirm full access to all data used in the study and accept responsibility for the journal submission.

## Data availability

The complete data set used for this study will be uploaded in a public repository upon acceptance.

## Ethics Approval, Role of Funding Sources

Ethics approval was provided by the Research Ethics Committee of the International Institute of Social Studies, Erasmus University Rotterdam.

The funding sources did not influence the design, interpretation of results and writing of the study.

## Appendix

**Table A1.** Provision of Covid-19 related health services.

| Type of service | Number of health facilities | March 2020 | April 2020 | May 2020 | June 2020 | March – June 2020 |
| --- | --- | --- | --- | --- | --- | --- |
| Quarantine | 21 | 17.6 | 76.1 | 171.8 | 544.1 | 202.4 |
| Testing | 42 | 68.8 | 106.4 | 398.8 | 369.4 | 235.8 |
| Outpatient Treatment | 18 | 17.2 | 46.8 | 103.4 | 118.2 | 71.4 |
| ICU bed | 9 | 0.8 | 3.1 | 5.4 | 8.7 | 4.5 |

**Note:** In total, 52 facilities provide Covid-19 related services.

**Table A2.** Infection Preventions Measures.

| No | Measures | % |
| --- | --- | --- |
| 1 | Daily temperature check for health workers when entering the facility | 92 |
| 2 | Temperature check for patients when entering the facility | 98 |
| 3 | Regularly disinfecting the health facility and medical equipment | 90 |
| 4 | Introducing social distancing measures for patients and health workers | 99 |
| 5 | Restricting entry to the facility for visitors and accompanying family members in order to reduce crowding in the facility | 91 |
| 6 | Mandatory use of safety equipment for health workers like gloves and masks | 97 |
| 7 | Mandatory use of safety equipment for patients like masks | 97 |
| 8 | Extending the duration of follow-up appointment | 84 |
| 9 | Offering telemedicine instead of in person follow-up visit | 36 |
| 10 | Reducing the number of days required for inpatient care | 60 |
| 11 | Reducing the number of medical staff per day | 52 |
| 12 | Reducing the number of administrative staff per day | 66 |
| 13 | Permitting stay at home for high-risk employees (those with underlying health conditions) or elder employees | 80 |

**Notes:** N is 87 or 88

**Table A3.**
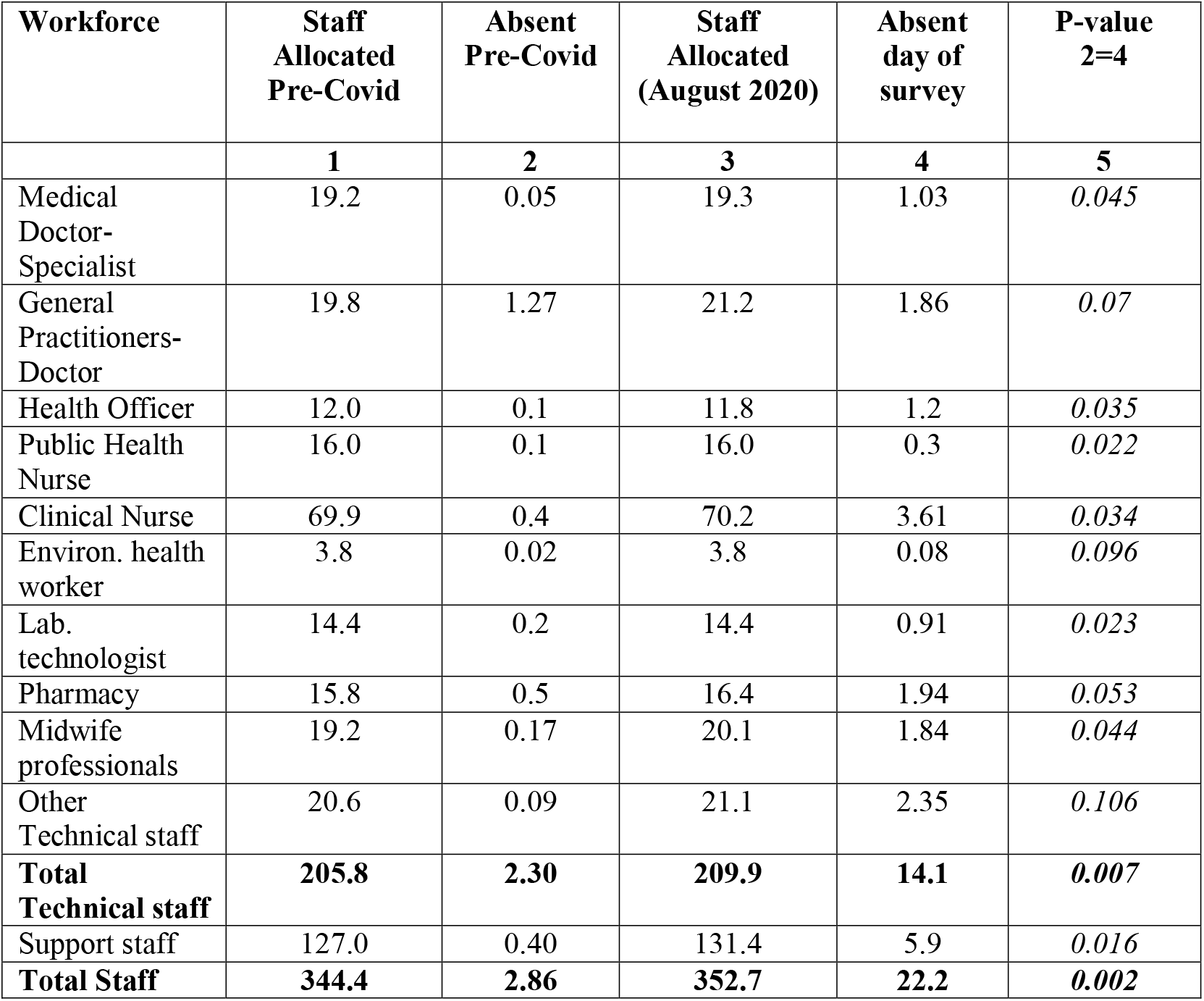
Absenteeism and Covid 19.

